# Graduate students significantly more concerned than undergraduates about returning to campus in the era of COVID-19

**DOI:** 10.1101/2020.07.15.20154682

**Authors:** Mark H. Ebell, Laura Bierema, Lauren Haines

## Abstract

**Introduction:** As students return to colleges and universities in the fall of 2020, it is important to understand their perception of risk and their desire for in person versus online learning, which may differ between undergraduate and graduate students.

**Methods:** We anonymously surveyed 212 undergraduate and 134 graduate students in the College of Public Health, and 94 graduate students in the College of Education in late June, 2020. We asked them Likert style questions regarding their comfort returning to campus and their preferred learning strategies once back. We compared “Strongly agree/Agree” with “Neutral/Disagree/Strongly disagree” using a chi-square test.

**Results:** Graduate students were significantly less likely to look forward to being on campus (38.3% doctoral vs 40.6% master’s vs 77.7% undergraduate, p < 0.001), more likely to perceive themselves as high risk (43.3% doctoral vs 40.0% masters vs 17.5% undergraduate, p < 0.001), and were more likely to prefer all classwork online (66.7% doctoral vs 44.6% master’s vs 20.8% undergraduate, p < 0.001). Graduate students were also less likely to prefer to be in the classroom as much as possible in the fall (59.2% doctoral vs 67.7% master’s vs 74.5% undergraduate, p < 0.001). Most were not concerned about their ability to conduct research. Students generally supported wearing of facemasks indoors.

**Conclusions:** There are important differences in perception of risk and desire for online versus in-person learning between undergraduate and graduate students. Faculty and administrators must acknowledge and address these differences as they prepare for return to campus in the fall.

## Introduction

The COVID-19 pandemic caused by the SARS-CoV-2 virus has created a worldwide health crisis. There have been over 10 million confirmed cases and over 500,000 deaths worldwide,^1^ with an estimated 10 undetected cases per confirmed case.^2^ As students return to campus in the fall, it is important to understand their concerns and willingness to engage in usual classroom activities. We hypothesize that undergraduate and graduate students may have different attitudes towards returning to campus based on differences in age, attitudes toward risk-taking behavior, likelihood of comorbid illness, and different living circumstances.^3,4^ Therefore, we anonymously surveyed students at the University of Georgia in the College of Public Health and the College of Education with regards to their attitudes toward returning to campus and their preferred learning environment.

## Methods

The purpose of this survey was initially to provide guidance to faculty and administrators in the College of Public Health at the University of Georgia regarding re-opening in the fall in the face of the COVID-19 pandemic. The survey was shared with faculty in the College of Education who had similar concerns. Once the survey results were examined, it was clear that they might be of interest to the larger academic community and the decision was made to publish the results. The survey was performed online in late June, 2020.

The questions used a Likert style format, with options of “Strongly disagree”, “Strongly agree”, “Neutral”, “Agree”, and “Strongly agree”. After surveing students in the Department of Epidemiology and Biostatistics, the survey was slightly adapted by leadership of the College of Public Health (the question “I am comfortable with a requirement that students and faculty wear masks in indoor spaces and during classes” was changed to “Strongly encouraging student to wear face masks while indoors or during face-to-face lectures is a good idea”, and the question “I prefer not to wear face masks while indoors or during face-to-face lectures” was added). Students in the other departments of the College of Public Health were then invited to participate, including graduate students and undergraduates. Finally, the survey was adapted by faculty in the program of Learning Leadership and Organizational Development in the College of Education and sent to their graduate students. They changed response categories from 5 to 4, eliminating “Neutral”.

For analysis, categories were collapsed to “Strongly agree or agree” versus “Neutral, disagree, or strongly disagree”. The chi-square test was used to evaluate statistical significance. Because the surveys were originally done to guide policy and not as research, the Human Subjects committee designated this as not human subjects research.

## Results

The survey was completed by 346 graduate and undergraduate students in the College of Public Health and 94 students in the College of Education. Among the College of Education students, all in the Department of Lifelong Education Administration and Planning, 79 were pursuing a doctoral degree and 15 a Master’s degree. Of the College of Public health students, 70 were pursuing a doctoral degree, 64 a Master’s degree, and 212 were undergraduates in Health Promotion and Behavior or Environmental Health Sciences.

Graduate students were significantly more cautious regarding the return to campus. They were less likely to agree with the statement that they were looking forward to being on campus (38.3% doctoral vs 40.6% master’s vs 77.7% undergraduate, p < 0.001), and more likely to perceive themselves as being high risk for serious illness (43.3% doctoral vs 40.0% masters vs 17.5% undergraduate, p < 0.001). They were also more likely to agree that they preferred to have all lectures and classwork online in the fall (66.7% doctoral vs 44.6% master’s vs 20.8% undergraduate, p < 0.001), and less likely to prefer to be in the classroom as much as possible in the fall (59.2% doctoral vs 67.7% master’s vs 74.5% undergraduate, p < 0.001). Just over one-third of public health students were concerned about their ability to conduct research and this did not differ by student category. Students generally supported wearing of facemasks while indoors.

Conversely, undergraduate students had the lowest percentages for agreeing they were concerned about acquiring COVID-19 upon return, for agreeing they wanted all classwork online this fall, and for agreeing they were apprehensive about coming back to campus. Responses of graduate students in the College of Education were compared with graduate students in the College of Public Health. Students in the College of Public Health were slightly less likely to favor a requirement to wear masks (74% vs 88%, p = 0.03). Otherwise there were no significant differences in responses to the other questions.

## Discussion

As students, faculty, staff, and administrators contemplate the return to campus this fall in the face of COVID-19, each group is assessing their comfort with in-person teaching and on campus meetings. In our survey, we found that students should not be treated as a monolithic group, and that graduate students were much more concerned about their risk of acquiring infection and significantly more comfortable with the idea of online instruction than undergraduates.

The disparity between undergraduate and graduate perspectives about on campus instruction and activities suggests that typical “Return to Campus Guidelines” do not cater to graduate student concerns. This survey is critical to understanding that diverse student groups have differing perspectives of risk. Given these range of perspectives regarding health and safety, universities should provide more latitude in how courses and meetings will be conducted in light of concerns for safety.

In 2018 there were approximately 1.8 million graduate students in the United States, of whom 57% were enrolled full-time with the largest enrollments in education, business, and health sciences.^5^ These fields also have the largest proportions of part-time graduate students who likely manage employment and dependent care in addition to graduate school. Notably, 57% of graduate students are women,^6^ who usually bear the burden of domestic tasks and dependent care. Further, research on homeschooling during the pandemic shows it is being handled disproportionately by women,^7^ and longitudinal data show working mothers shoulder the bulk of childcare and housework without reductions in work hours of more than 2 hours per day as compared to 40 minutes for men.^8^

The average age of graduate students in the United States is approximately 33 years old,^4^ a figure that has remained static over the years, with 22% of all graduate students over the age of 40 years.^9^ Only 48% of graduate students are unmarried with no dependents,^4^ suggesting that over half of graduate students tend to have more adult-like responsibilities than undergraduates such as employment and dependent care, potentially putting graduate students at higher risk of exposing family members and work colleagues to COVID-19 if they are attending courses on campus.

## Conclusions

It is important for university programs to take an evidence-based and flexible approach to the return to campus, one that addresses the differing concerns of graduate and undergraduate students. This is especially important in health and leadership professions where students are learning how critical decisions affect the health and wellbeing of organizations and communities. Administrators are advocating a “HyFlex” approach to instruction. We advocate that university administrators should adopt a similarly flexible approach that emboldens faculty and students to make decisions about pedagogy that reflect their personal risk factors, concerns, and goals.

**Table 1.**
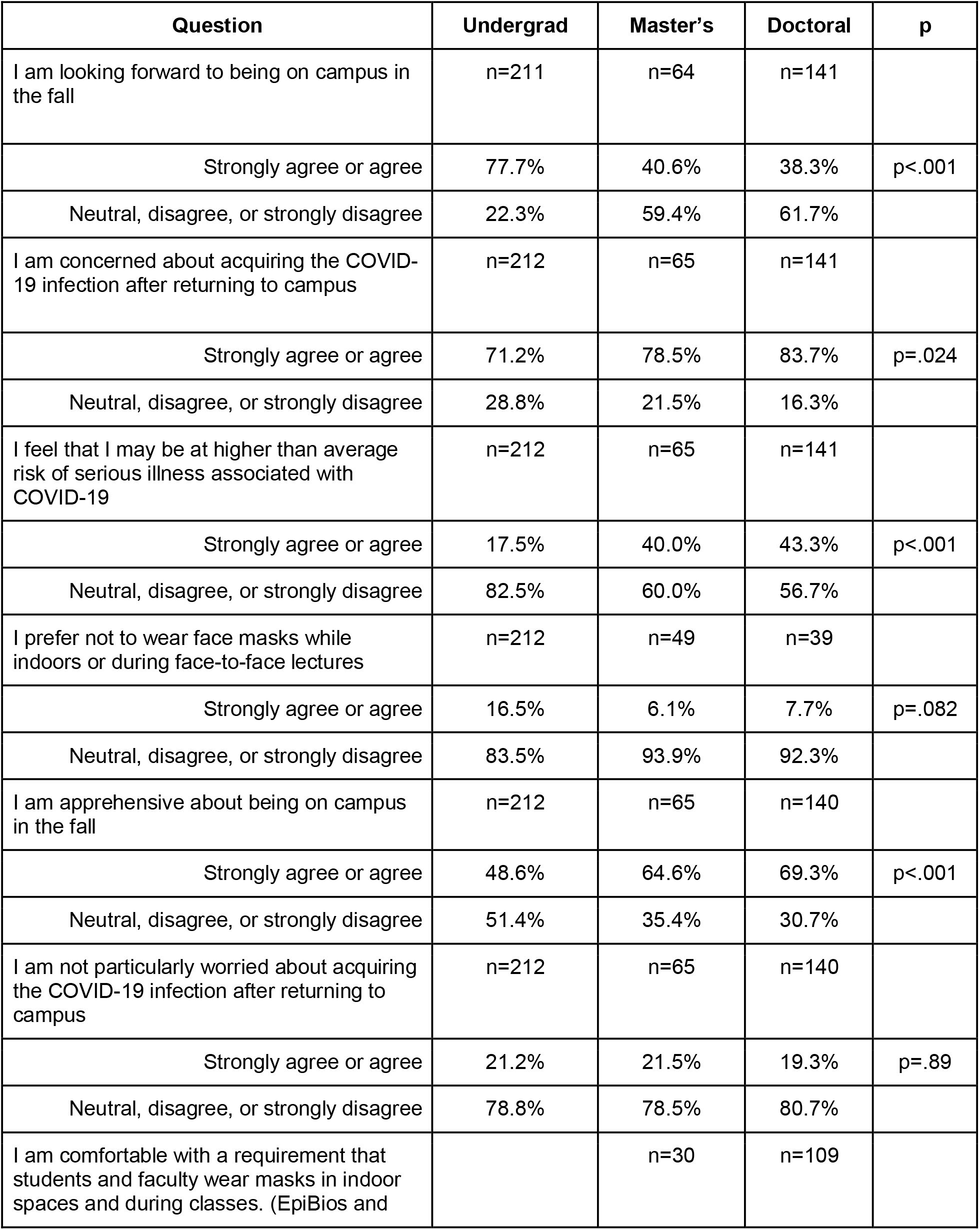
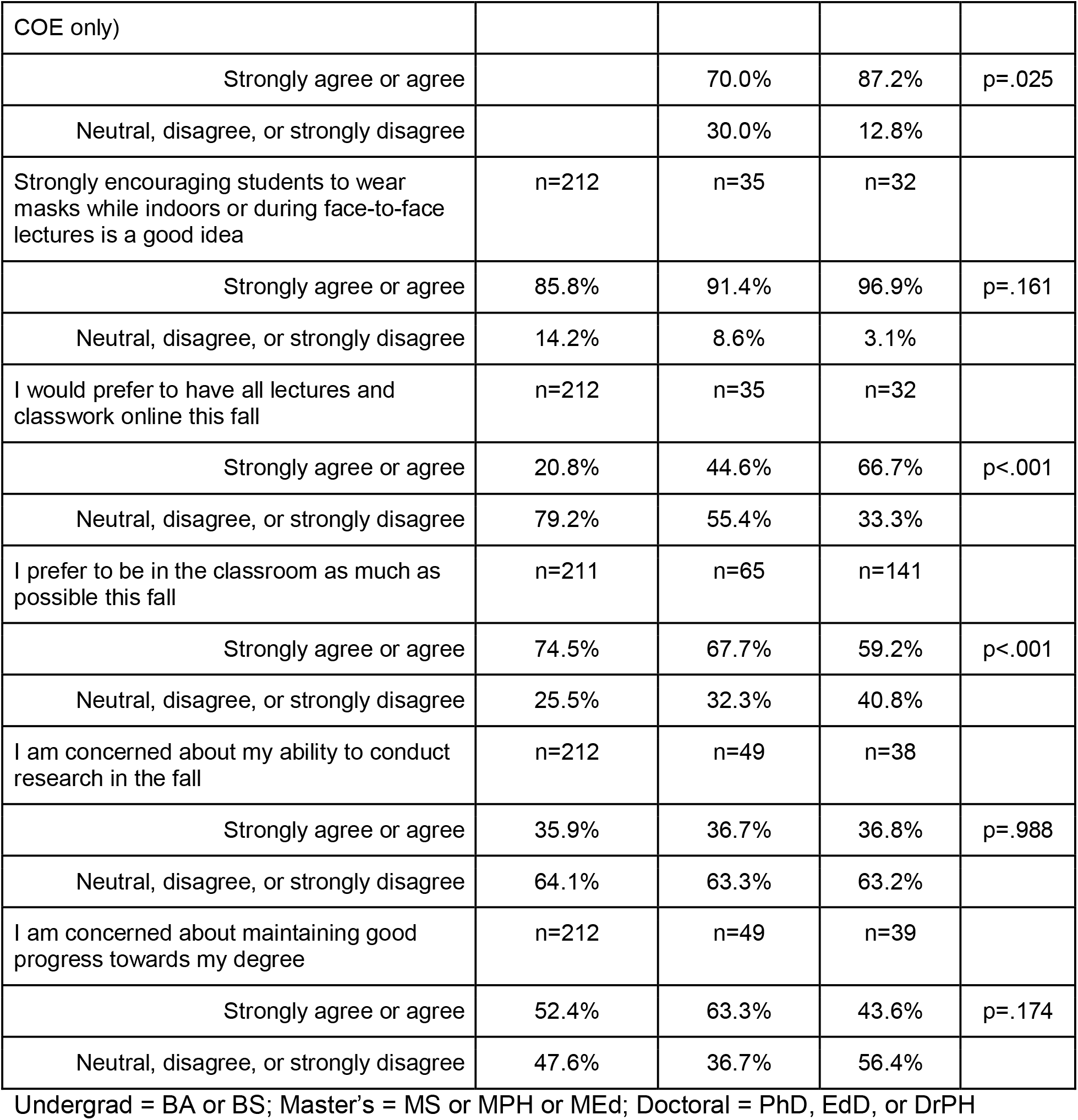
were less likely to want to be on campus

| Question | Undergrad | Master's | Doctoral | p |
| --- | --- | --- | --- | --- |
| I am looking forward to being on campus in the fall | n=211 | n=64 | n=141 |  |
| Strongly agree or agree | 77.7% | 40.6% | 38.3% | p<.001 |
| Neutral, disagree, or strongly disagree | 22.3% | 59.4% | 61.7% |  |
| I am concerned about acquiring the COVID-19 infection after returning to campus | n=212 | n=65 | n=141 |  |
| Strongly agree or agree | 71.2% | 78.5% | 83.7% | p=.024 |
| Neutral, disagree, or strongly disagree | 28.8% | 21.5% | 16.3% |  |
| I feel that I may be at higher than average risk of serious illness associated with COVID-19 | n=212 | n=65 | n=141 |  |
| Strongly agree or agree | 17.5% | 40.0% | 43.3% | p<.001 |
| Neutral, disagree, or strongly disagree | 82.5% | 60.0% | 56.7% |  |
| I prefer not to wear face masks while indoors or during face-to-face lectures | n=212 | n=49 | n=39 |  |
| Strongly agree or agree | 16.5% | 6.1% | 7.7% | p=.082 |
| Neutral, disagree, or strongly disagree | 83.5% | 93.9% | 92.3% |  |
| I am apprehensive about being on campus in the fall | n=212 | n=65 | n=140 |  |
| Strongly agree or agree | 48.6% | 64.6% | 69.3% | p<.001 |
| Neutral, disagree, or strongly disagree | 51.4% | 35.4% | 30.7% |  |
| I am not particularly worried about acquiring the COVID-19 infection after returning to campus | n=212 | n=65 | n=140 |  |
| Strongly agree or agree | 21.2% | 21.5% | 19.3% | p=.89 |
| Neutral, disagree, or strongly disagree | 78.8% | 78.5% | 80.7% |  |
| I am comfortable with a requirement that students and faculty wear masks in indoor spaces and during classes. (EpiBios and |  | n=30 | n=109 |  |
| COE only) |  |  |  |  |
| Strongly agree or agree |  | 70.0% | 87.2% | p=.025 |
| Neutral, disagree, or strongly disagree |  | 30.0% | 12.8% |  |
| Strongly encouraging students to wear masks while indoors or during face-to-face lectures is a good idea | n=212 | n=35 | n=32 |  |
| Strongly agree or agree | 85.8% | 91.4% | 96.9% | p=.161 |
| Neutral, disagree, or strongly disagree | 14.2% | 8.6% | 3.1% |  |
| I would prefer to have all lectures and classwork online this fall | n=212 | n=35 | n=32 |  |
| Strongly agree or agree | 20.8% | 44.6% | 66.7% | p<.001 |
| Neutral, disagree, or strongly disagree | 79.2% | 55.4% | 33.3% |  |
| I prefer to be in the classroom as much as possible this fall | n=211 | n=65 | n=141 |  |
| Strongly agree or agree | 74.5% | 67.7% | 59.2% | p<.001 |
| Neutral, disagree, or strongly disagree | 25.5% | 32.3% | 40.8% |  |
| I am concerned about my ability to conduct research in the fall | n=212 | n=49 | n=38 |  |
| Strongly agree or agree | 35.9% | 36.7% | 36.8% | p=.988 |
| Neutral, disagree, or strongly disagree | 64.1% | 63.3% | 63.2% |  |
| I am concerned about maintaining good progress towards my degree | n=212 | n=49 | n=39 |  |
| Strongly agree or agree | 52.4% | 63.3% | 43.6% | p=.174 |
| Neutral, disagree, or strongly disagree | 47.6% | 36.7% | 56.4% |  |

Undergrad = BA or BS; Master's = MS or MPH or MEd; Doctoral = PhD, EdD, or DrPH

## Data Availability

The investigators will make the original data available to other researchers upon request.

## Acknowledgements

The authors thank the students in the University of Georgia’s Colleges of Public Health and Education for their time spent completing this survey.

## References

1. Johns Hopkins COVID-19 Dashboard. https://coronavirus.jhu.edu/map.html. Accessed June 27, 2020.

2. Stringhini S, Wisniak A, Piumatti G, et al. Seroprevalence of anti-SARS-CoV-2 IgG antibodies in Geneva, Switzerland (SEROCoV-POP): a population-based study. Lancet. 2020. doi:10.1016/s0140-6736(20)31304-0

3. Romer D. Adolescent risk taking, impulsivity, and brain development: Implications for prevention. Dev Psychobiol. 2010. doi:10.1002/dev.20442

4. NCES. National Postsecondary Student Aid Study National Postsecondary Student Aid Study (NPSAS). In: NCES HANDBOOK OF SURVEY METHODS.; 2017.

5. Okahana H, Zhou E. Graduate Enrollment and Degrees: 2007-2017. Washington, DC Counc Grad Sch. 2018.

6. Choy SP, Cataldi EF, National Center for Education Statistics (ED) DC. W, MPR Associates CA. B. Student Financing of Graduate and First-Professional Education: 2003-04. Profiles of Students in Selected Degree Programs and Part-Time Students. Statistical Analysis Report. NCES 2006-185.; 2006.

7. Miller CC. Nearly Half of Men Say They Do Most of the Home Schooling. 3 Percent of Women Agree. The New York Times. https://www.nytimes.com/2020/05/06/upshot/pandemic-chores-homeschooling-gender.html.

8. Yavorsky JE, Kamp Dush CM, Schoppe-Sullivan SJ. The Production of Inequality: The Gender Division of Labor Across the Transition to Parenthood. J Marriage Fam. 2015. doi:10.1111/jomf.12189

9. Bell NE. Council of Graduate Schools (2009). Research report: Data Sources: Non-Traditional Students in Graduate Education. https://cgsnet.org/ckfinder/userfiles/files/DataSources_2009_12.pdf. Accessed July 14, 2020.

